# Evaluation of statistical detection of change algorithm for triaging multiple sclerosis patients with new lesion activity on longitudinal brain MRI

**DOI:** 10.1101/2023.01.31.23285297

**Authors:** Moayad Homssi, Elizabeth M. Sweeney, Emily Demmon, William Mannheim, Michael Sakirsky, Yi Wang, Susan A. Gauthier, Ajay Gupta, Thanh D. Nguyen

**Affiliations:** Department of Radiology, Weill Cornell Medicine, New York, NY, USA; Penn Statistics in Imaging and Visualization Endeavor (PennSIVE) Center, Department of Biostatistics, Epidemiology, and Informatics, University of Pennsylvania, Philadelphia, PA, USA; Department of Neurology, Weill Cornell Medicine, New York, NY, USA; The Feil Family Brain & Mind Institute, Weill Cornell Medicine, New York, NY, USA

## Abstract

**Background and Purpose:** Identification of new MS lesions on longitudinal MRI by human readers is time-consuming and prone to error. Our objective was to evaluate the improvement in a subject-level detection performance by readers when assisted by the automated statistical detection of change (SDC) algorithm.

**Materials and Methods:** A total of 200 MS patients with mean inter-scan interval of 13.2 ± 2.4 months were included. SDC was applied to the baseline and follow-up FLAIR images to detect potential new lesions for confirmation by readers (Reader+SDC method). This method was compared with readers operating in the clinical workflow (Reader method) for a subject-level detection of new lesions.

**Results:** Reader+SDC found 30 subjects (15.0%) with at least one new lesion, while Reader detected 16 subjects (8.0%). As a subject-level triage tool, SDC achieved a perfect sensitivity of 1.00 (95% CI: [0.88, 1.00]) and a moderate specificity of 0.67 (95% CI: [0.59, 0.74]). The agreement on a subject-level was 0.91 (95% CI: [0.87, 0.95]) between Reader+SDC and Reader, and 0.72 (95% CI: [0.66, 0.78]) between Reader+SDC and SDC.

**Conclusion:** SDC improves the detection accuracy of human readers and can serve as a time-saving patient triage tool for detecting new MS lesion activity on longitudinal FLAIR images.

## Introduction

Detection of new lesion activity on serial MRI is important for the disease diagnosis, monitoring, treatment response evaluation in MS patients.^1^ In most clinical workflows, expert readers manually view baseline and follow-up brain MRI images side-by-side on a PACS monitor to look for voxels with sufficiently large change in image intensity and size to be considered a potentially clinically relevant new MS lesion.^2^ The state-of-the-art 3D T2W FLAIR images acquired in a routine clinical MS imaging protocol^1^ provide high 1 mm isotropic resolution and excellent soft tissue contrast for lesion detection. However, native images obtained at two different time points are often imperfectly aligned due to differences in patient positioning and acquisition technique. Therefore, detecting new lesions by visual matching on the unregistered longitudinal images in the presence of noise is a time-consuming, error-prone, and highly observer-dependent task, even for human experts.^3^

A number of automated and semi-automated algorithms have been developed to overcome these challenges.^4-6^ In the classical approach, serially acquired images are intensity-normalized and co-registered, from which a dissimilarity map (e.g., obtained by subtraction) is calculated and then automatically segmented (e.g., by thresholding or statistical inference methods) or reviewed by humans to yield the final lesion change mask.^2, 7-14^ More recently, supervised deep learning-based convolutional neural network models have become the predominant approach.^15-19^ Despite rapid advances in research, the detection sensitivity and specificity remain moderate on a voxel- or lesion-level (less than 0.8).^3, 6^ We previously introduced the statistical detection of change algorithm (SDC) algorithm as an automated lesion change detection tool to visually assist human readers. This algorithm applies an optimal binary change detector to the subtraction of two longitudinally registered FLAIR images to delineate brain areas with potential new lesions.^13^ The purpose of this study was to evaluate the improvement in a subject-level detection performance by human readers when assisted by SDC, in comparison with the benchmark of human readers operating in the clinical workflow.

## METHODS

### Study cohort

This was a retrospective longitudinal study conducted in a cohort of 200 MS patients (145 women (72.5%), 55 men (27.5%); mean age, 47.6 years ± 10.9 (standard deviation [SD]); range, 18.5-75.8 years) who were enrolled in an ongoing prospective imaging and clinical database for MS research. The database was approved by the local Institutional Review Board and written informed consent was obtained from all participants prior to their entry into the database. Consecutive patients who underwent two MRI scans between September 20, 2017, and July 7, 2021, with a mean follow-up interval of 13.2 ± 2.4 months (range, 7.5-24.8 months) were included. The final cohort consisted of 6 patients with clinically isolated syndrome, 181 with relapsing-remitting MS, 6 with primary progressive MS, and 7 with secondary progressive MS. The mean disease duration was 14.7 years ± 7.4 (range, 2.6-54.9 years) and the mean Expanded Disability Status Scale (EDSS) was 1.3 ± 1.6 (range, 0.0-7.0; median, 1.0; interquartile range, 2.0).

### MRI examinations

Patients were imaged on Siemens 3T MRI scanners (Siemens Healthineers, Erlangen, Germany) using a product 20-channel head/neck coil. The scanning protocol contained 3D T1W MPRAGE sequence for anatomical definition and 3D T2W FLAIR sampling perfection with application optimized contrasts using different flip angle evolution (SPACE) sequence for lesion identification, using the following imaging parameters: 1) 3D sagittal T1W MPRAGE: TR/TE/TI = 2300.0/2.3/900 ms, flip angle (FA) = 8°, bandwidth (BW) = 200 Hz/pixel, acquired voxel size = 1.0 mm isotropic, number of slices = 176, parallel imaging factor (R) = 2.0, scan time = 5:21 min; 2) 3D sagittal T2W FLAIR SPACE: TR/TE/TI = 7600/448/2450 ms, FA = 90°, BW = 781 Hz/pixel, echo spacing = 3.42 ms, turbo factor = 284, acquired voxel size = 1.0 mm isotropic, number of slices = 176, R = 4.0, scan time = 5:21 min.

### Image post-processing

At each timepoint, T1W and FLAIR images were brain extracted using FMRIB Software Library (FSL) BET command,^20^ and corrected for spatial inhomogeneity and segmented into gray matter, white matter, and CSF masks using FSL FAST command.^21^ The FLAIR image was then linearly registered to the T1W structural image using FSL FLIRT command^22^ with 6 degrees of freedom (rigid body transformation). For longitudinal registration, the baseline and follow-up brain-extracted T1W images were first linearly aligned to a halfway space^8^ using ANTs algorithm with 12 degrees of freedom (rigid body and affine transformation),^23^ followed by registration of the corresponding FLAIR images into the same halfway space using the concatenated transformation matrices obtained from the previous steps. The purpose of spatially aligning longitudinal images to the halfway space was to ensure that the degree of blurring introduced by the registration algorithm was similar between images, which improves image subtraction. Next, the SDC algorithm,^13^ implemented in MATLAB R2020 (MathWorks, Natick, MA, USA), was applied to the registered FLAIR images in the halfway space to detect brain voxels with positive signal change (indicating new lesions or growth of existing lesions). Finally, the detected changed voxels were registered back to the follow-up FLAIR image. To facilitate subsequent visual confirmation of the detected new lesions by human readers, a binary mask was generated by the SDC algorithm which was overlaid on the baseline and follow-up FLAIR image pair to delineate potential new lesions with a red box (Fig.1).

**Figure 1.**
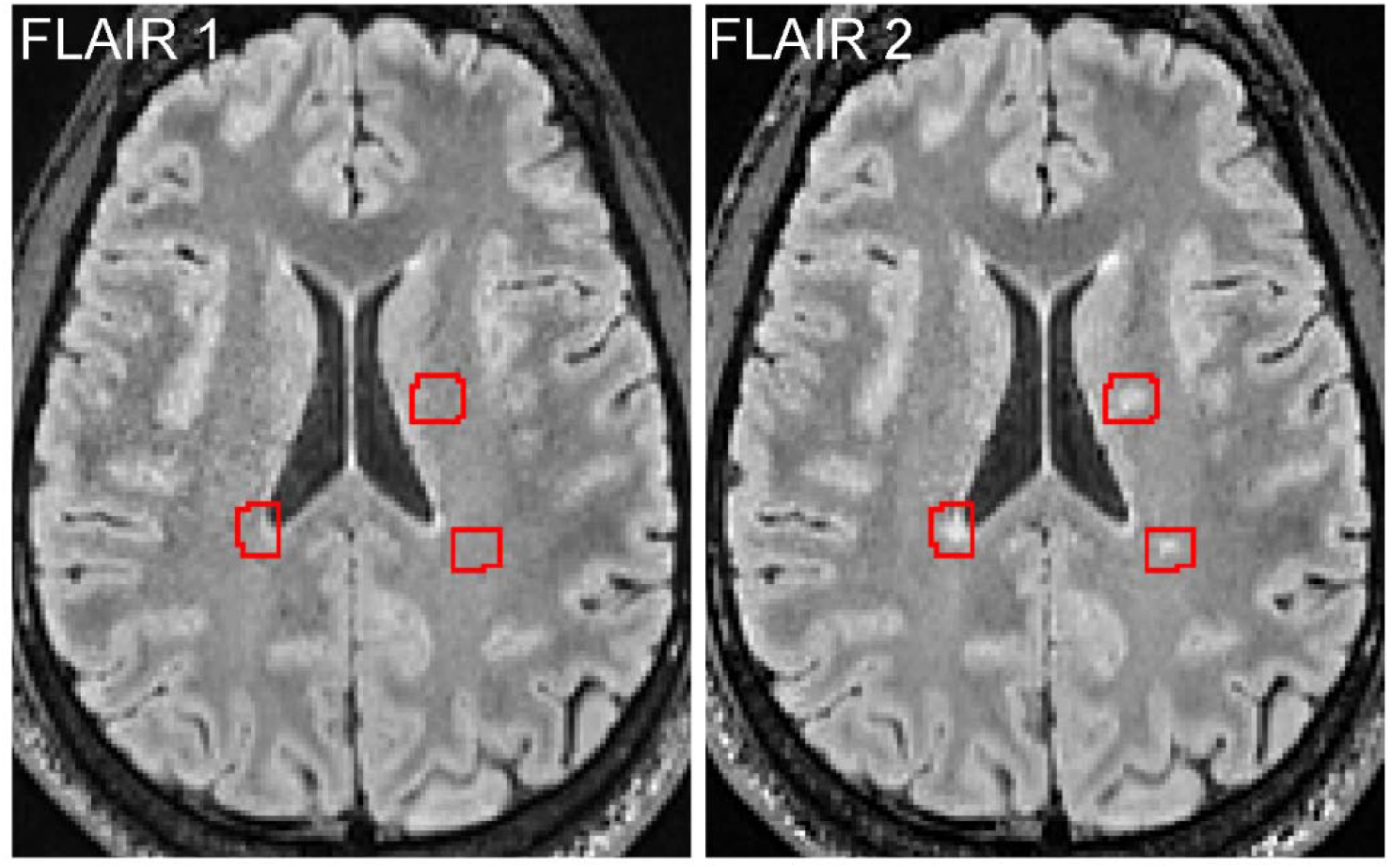
Longitudinal axial FLAIR images acquired from an MS patient approximately 10.5 months apart, showing an example of three new lesions detected by human readers who were assisted visually by the SDC algorithm which delineates brain areas containing potential new lesions for human confirmation (referred to as Reader+SDC method in the text).

### Visual identification of new lesions

The registered baseline and follow-up FLAIR images, along with the red boxes marking the potential new lesions detected by SDC (Fig.1), were displayed side-by-side in the axial plane using ITK-SNAP v3.8 software.^24^ Two expert readers⎯a board-certified neuroradiologist with 16 years of experience and an MR physicist with 20 years of experience, both of whom were blinded to clinical and other imaging information⎯jointly reviewed the FLAIR images with visual assistance from SDC-detected areas of lesion growth to identify new lesions based on consensus (Reader+SDC method). A lesion was considered as new if it could be seen on the follow-up FLAIR image but could not be ascertained on the baseline FLAIR image. To assess the influence of lesion size on new lesion detection performance, the binary mask generated by SDC was manually checked and edited if necessary for each confirmed new lesion and the lesion volume was recorded.

For comparison, the official radiology reports created by board-certified neuroradiologists at the time of the clinical encounters were retrieved from EPIC electronic medical record system (Epic Systems Corporation, Verona, WI, USA) and manually parsed for the mention of at least one new lesion. In our routine clinical workflow, new lesions are detected based on the visual inspection of baseline and follow-up native FLAIR images as well as T1W, T2W and Gadolinium-enhanced T1W images (hereafter referred to as Reader method). The total number of new lesions and their precise anatomical locations are variably recorded in the reports subject to the clinical scenario and preferences of the interpreting radiologist. Therefore, in this study, lesion detection outcome for the three methods (Reader, SDC, and Reader+SDC) was defined at a subject-level as a binary indicator of having at least one new lesion.

### Statistical analysis

All statistical analysis was performed in R v4.1.2.^25^ We were interested in the detection of one or more new lesions at a subject-level. Change in size of existing lesions (growth or shrinkage) and lesions smaller than 15mm^3^ were excluded from the analysis. This lesion size cutoff was calculated assuming a spherical lesion shape with a diameter of 3 mm (3 voxels on our FLAIR image) in accordance with the currently accepted minimum lesion size on MRI.^1^ For each subject and each method (Reader, SDC, and Reader+SDC), a binary indicator of incidence of new lesions was created. Contingency tables at a subject-level were investigated and sensitivity, specificity, and positive predictive value (PPV) were assessed using the Reader+SDC method as a reference. Exact binomial 95% confidence intervals were calculated for these measures.^26^ Agreement between all the methods was calculated with non-parametric bootstrapped 95% confidence intervals.^27^

In addition, analysis of the effect of the new lesion size on the detection performance was performed in those subjects who were determined to have new lesion activity by Reader+SDC method. We tested if there were differences in the size of individual new lesions identified by both Reader and Reader+SDC methods versus those identified by Reader+SDC method but not by Reader method. A linear mixed-effects model was fit with the log of the individual lesion volume as an outcome regressed on an indicator of whether Reader and Reader+SDC methods were in agreement at a subject-level. A random effect for patient was included in the model to account for the correlation in the data resulting from multiple new lesions per subject. In addition, the log of the total new lesion volume for each patient and the total number of new lesions was calculated in patients where new lesion activity was found by Reader+SDC. A t-test was performed to assess if there were differences between subjects where Reader+SDC and SDC agreed and disagreed. A p-value of less than 0.05 was considered statistically significant.

## RESULTS

All 200 pairs of baseline and follow-up FLAIR scans were interpretable. Of these, 80 pairs (40.0%) were acquired on the same MRI scanner. The fully automated SDC algorithm detected 86 subjects (43.0%) with at least one potential new lesion. The semi-automated Reader+SDC method, in which two readers identified new lesions by comparing the two longitudinally registered FLAIR images with visual assistance from SDC showing potential new lesions (Fig.1), detected 41 subjects (20.5%) with at least one new lesion. After excluding lesions smaller than 15 mm^3^, Reader+SDC found 30 individuals (15.0%) with new lesion(s). In comparison, the traditional Reader method, performed by radiologists in the routine clinical workflow, identified 20 subjects (10.0%) with at least one new lesion.

Table 1 shows the contingency table for subject-level detection of new lesions on FLAIR images obtained by the Reader and SDC methods using the Reader+SDC method as a reference (note that lesions smaller than 15 mm^3^ were excluded from the statistical analysis for SDC and Reader+SDC methods, while the Reader method did not provide lesion size information). Over 200 MS cases, Reader failed to detect new lesions in 14/30 individuals, resulting in a moderate sensitivity of 0.53 (95% CI: [0.34, 0.72]), with an excellent specificity of 0.98 (95% CI: [0.94, 0.99]) and a good PPV of 0.80 (95% CI: [0.56, 0.94]). The four subjects that were identified as having at least one new lesion by Reader but not by Reader+SDC in Table 1 were all found to have new lesions smaller than the 15 mm^3^ cutoff. In comparison, SDC was able to detect all 30/30 patients with at least one new lesion, achieving a perfect sensitivity of 1.00 (95% CI: [0.88, 1.00]), although at the cost of lower specificity of 0.67 (95% CI: [0.59, 0.74]) and lower PPV of 0.35 (95% CI: [0.25, 0.46]). The agreement on a subject-level was found to be 0.91 (95% CI: [0.87, 0.95]) between Reader+SDC and Reader, 0.72 (95% CI: [0.66, 0.78]) between Reader+SDC and SDC, and 0.64 (95% CI: [0.57, 0.71]) between Reader and SDC.

**Table 1.** Contingency table comparing a subject-level detection of new lesions obtained from longitudinal FLAIR images of 200 MS patients using the Reader (manual), SDC (fully automated) and Reader+SDC (semi-automated) methods. Reader+SDC was considered a reference method to provide the ground truth for comparison.

|  |  | Reader+SDC |  | Total |
| --- | --- | --- | --- | --- |
|  |  | New lesions | No new lesions |  |
| Reader | New lesions | 16 | 4 | 20 |
|  | No new lesions | 14 | 166 | 180 |
| SDC | New lesions | 30 | 56 | 86 |
|  | No new lesions | 0 | 114 | 114 |
| Total |  | 30 | 170 | 200 |

Figure 2 shows examples of four new lesions of various size and location from four different MS subjects that were identified by SDC and confirmed by Reader+SDC but were not detected by Reader according to the radiology report. Figure 3 shows examples of two new punctate lesions that were identified by Reader in the clinical workflow but were not detected by SDC and Reader+SDC after applying a minimum lesion volume threshold of 15 mm^3^.

**Figure 2.**
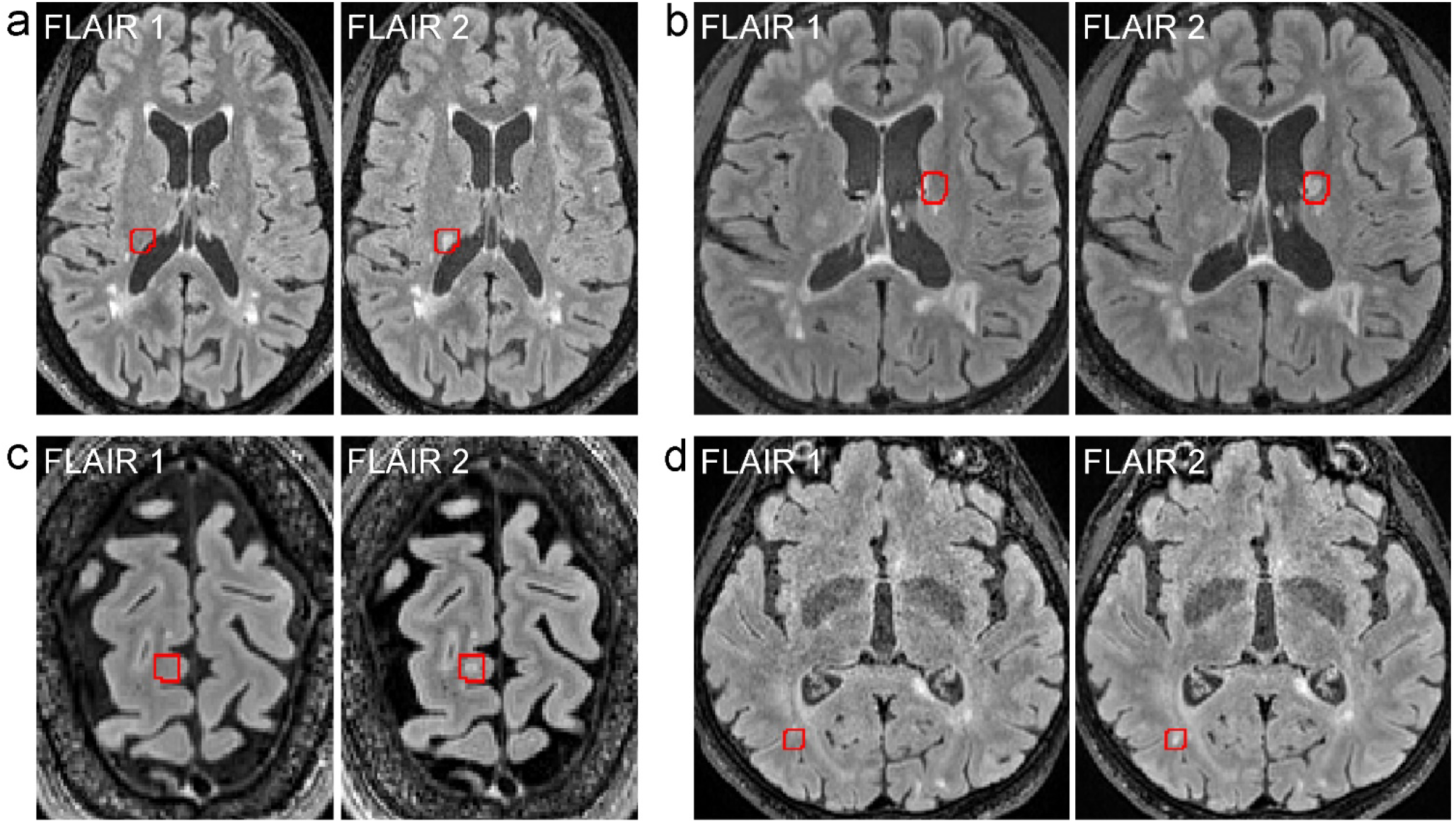
Examples of new lesions varying in size and location on longitudinal FLAIR images obtained from four different MS subjects. These lesions were initially identified by SDC (marked by red boxes) and later confirmed by Reader+SDC method but were not detected by Reader method using the conventional radiology workflow.

**Figure 3.**
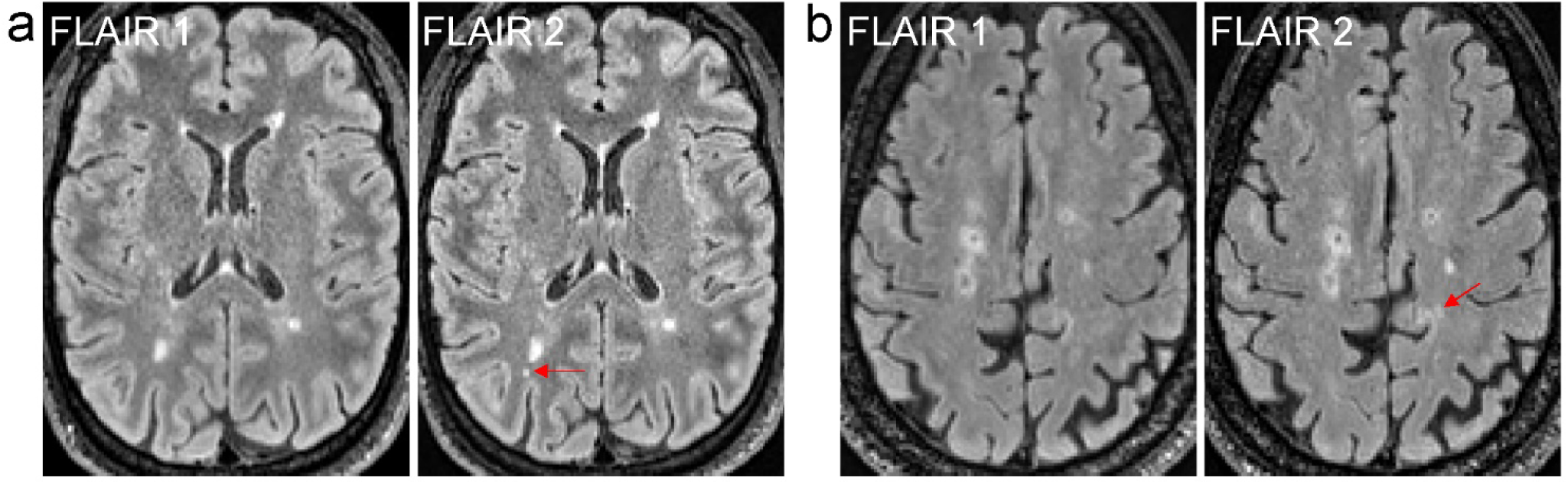
Examples of new punctate lesions on longitudinal FLAIR images acquired from two different MS subjects that were identified by Reader method in the clinical workflow but were not detected by SDC and Reader+SDC methods after applying a minimum lesion volume cutoff of 15 mm^3^.

For the lesion volume analysis, in the linear mixed effects model, no difference in the individual log new lesion volume was found between subjects identified by both Reader+SDC and Reader as having a new lesion (agreement group) and those identified by Reader+SDC but not by Reader (disagreement group) (p-value = 0.787). On a subject-level, a difference in the total log volume of new lesions was found (p-value = 0.013), with the total log lesion volume being on average smaller in the disagreement group. Figure 4 shows the boxplots of the total log lesion volume for subjects in the agreement and disagreement groups. The average log lesion volume in the agreement group was 5.72 versus 4.42 in the disagreement group. On average, the number of new lesions in patients with agreement between Reader+SDC and Reader was higher than that in those with disagreement (4.43 versus 1.24, p-value = 0.007).

**Figure 4.**
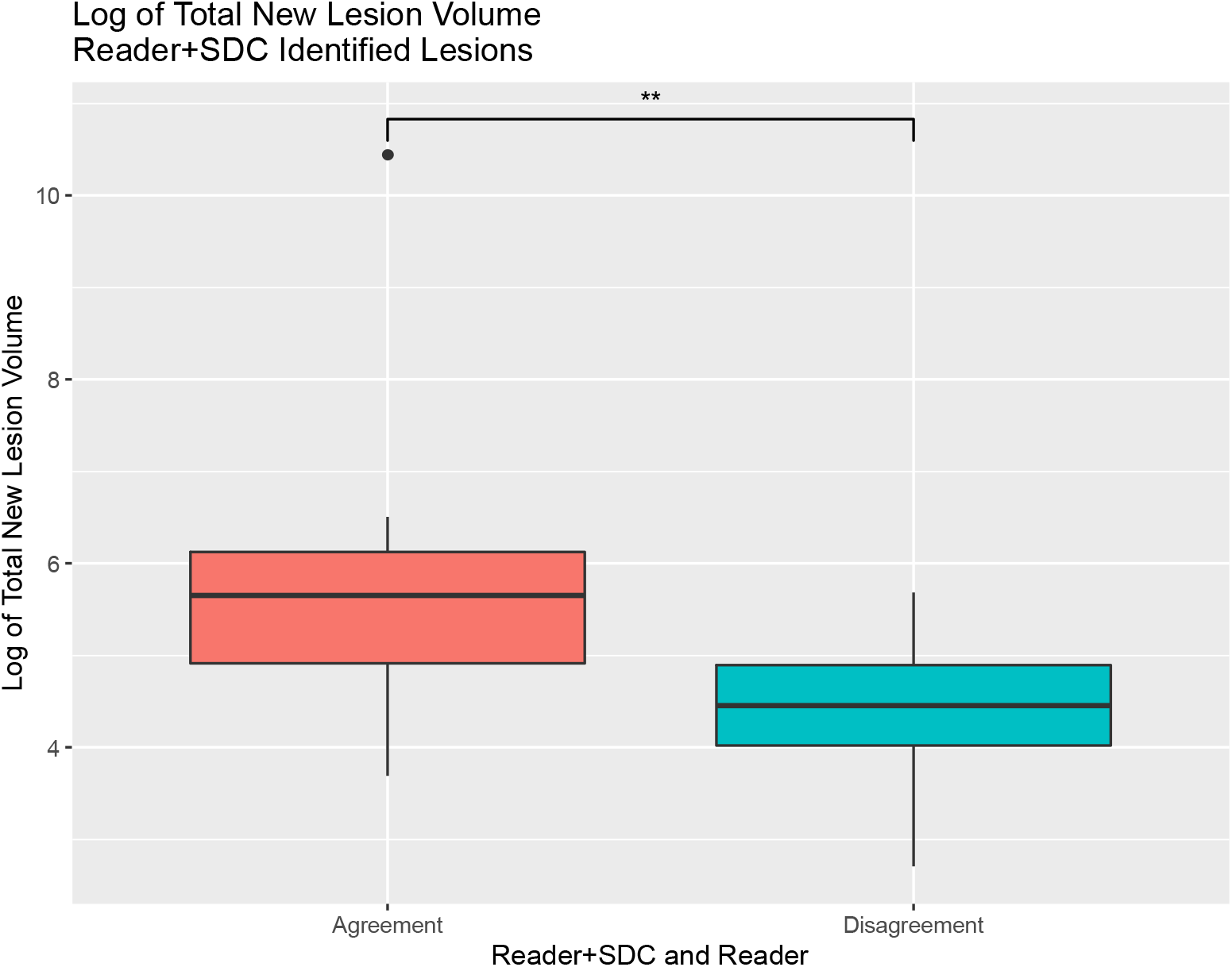
Boxplots of the log of total new lesion volume for MS subjects with new lesions identified by both Reader+SDC and Reader methods (agreement) compared to those with new lesions detected by Reader+SDC but not by Reader (disagreement). The log total new lesion volume was on average smaller in the disagreement group (p-value = 0.013).

## DISCUSSION

In this single-center study evaluating the utility of the automated SDC algorithm in assisting human readers to detect new lesions on longitudinal FLAIR images, we found that SDC was able to provide a perfect new lesion detection sensitivity on a subject-level, which represents an approximately two-fold improvement over the sensitivity of 0.53 of the human reader alone. SDC achieved this excellent level of detection sensitivity while providing a moderate subject-level specificity of 0.67 (meaning about two out of every three subjects without new lesion activity was correctly classified). These operating characteristics allow SDC to be used as a valuable triage tool and could accelerate the interpretation time of cases in which no new lesions are identified by SDC. In our study, for example, 114 of 200 patients had no new lesions on follow-up confirmed by SDC, suggesting that a more rapid expert human review of these cases (57% of our entire cohort) may be feasible. Such increase in efficiency may enable expert human readers to allocate more time to interpret MRI cases flagged by SDC as being potentially positive for new lesions.

In the conventional radiology workflow, detecting new lesions that formed between two longitudinal scans of MS patients is often performed by radiologists and other clinicians manually on a PACS monitor by comparing a large number of paired but imperfectly aligned FLAIR images. This approach is often time-consuming, mentally demanding, and error-prone, especially if there is a substantial image misalignment due to the difference in head orientation between the two scans. SDC overcomes these challenges by providing an automated detection of potential new lesions which are then visually indicated to the reader on a pair of longitudinally registered images (Fig.1). Conceptually, SDC is formulated as an optimal change detector applied to the subtraction image, which can be proved mathematically by the Neyman-Pearson lemma to provide the best detection power for a given false positive rate.^28^ By design, SDC mimics a human reader in two key aspects. First, it applies an adaptive intensity threshold to the subtraction image based on the level of noise in the image. For example, the threshold for the longitudinal signal change is automatically increased by SDC for more noisy images, which can be regarded as equivalent to setting a higher level of trust as often done by a human reader when dealing with noise. Second, SDC utilizes signal from voxels in a local neighborhood to calculate the test statistic, which helps to increase the detection reliability. Similar to change detection by humans, spatially spurious signals on the subtraction image are encoded by the algorithm to have lower likelihood of being identified as true change. By virtue of being capable to operate at a very high sensitivity and a reasonable specificity, SDC enables the new lesion identification problem to be shifted from the traditional difficult task of locating new lesions on unmarked images to a much easier task of confirming true positives and eliminating false positives in areas already marked out by SDC. Therefore, SDC has a great potential to shorten new lesion detection time while reducing reader fatigue.

There have been several studies demonstrating the benefits of detecting new lesions on a subject-level on longitudinally registered or subtracted standard 1 mm isotropic 3D FLAIR images. A study by Galletto et al performed in 94 MS patients showed that using an automated coregistration-fusion method improved the detection rate of subjects with at least one new lesion from 46% to 59%.^14^ In another study conducted by Echinger et al in 106 MS patients, 58% of subjects were identified as having at least one new lesion on the FLAIR subtraction image, with a similar proportion (59%) identified using the conventional reading method.^12^ In this study of 200 MS patients, we saw an improvement from 8% by Reader to 15% by Reader+SDC in subject-level detection rate, using a minimum lesion volume threshold of 15 mm^3^. The observed relatively rare event of a new lesion occurring in our cohort is likely related to different patient characteristics.

In this study, we did not find any difference in the volume of individual new lesions in subjects that Reader+SDC and Reader methods agreed and disagreed upon on a subject-level. However, we did find on average higher new lesion volume and a larger number of individual lesions in the agreement group. This is intuitive, as subjects with a higher new lesion volume and higher number of new lesions will be more likely to be recognized as having new lesion activity. These results indicate that using SDC may help in cases that would otherwise be missed due to low new lesion volume or a small number of new lesions.

This study has several limitations. First, cortical GM lesions were not considered due to the insufficient resolution of a routine clinical 3T MRI protocol. Second, in the current study design, the Reader method was performed in the routine clinical workflow by readers different from those involved in the Reader+SDC method. These differences make it difficult to determine the clinical significance, if any, of discrepancies in new lesion identification noted between the clinical radiologic report and retrospectively performed SDC-enhanced readings. For example, the radiologist issuing the clinical report in the Reader method had real-time access to the electronic health record and contrast-enhanced imaging while being subject to time constraints imposed in the clinical workflow and being responsible for total brain MRI interpretation, not only new lesion detection on FLAIR. While our study design allows a direct comparison of SDC with the radiology report, regarded as a clinically established benchmark, future work will be focused on integrating the SDC algorithm into the clinical workflow, which will allow for prospective comparison studies to be performed in a real-world setting. Third, a minimum lesion volume cutoff of 15 mm^3^ was used to mitigate the effect of noise on the false positive rate of SDC algorithm. Such punctate MS lesions are often considered inconsequential,^1^ although further evidence on their role in the disease progression and outcome may be needed. Finally, linear longitudinal brain registration was used in this study, which was deemed sufficiently accurate for the annual follow-up interval in our cohort but may not capture non-linear changes in the brain morphology over a longer inter-scan period. The use of deformable motion model in the registration algorithm may be considered in such scenario.^11, 23^

## CONCLUSIONS

The SDC algorithm improves the detection accuracy of human readers and can serve as a time-saving patient triage tool for detecting new MS lesion activity on longitudinal FLAIR images.

## Data Availability

All data produced in the present study are available upon reasonable request to the authors.

## Abbreviations

SDC: statistical detection of change
SPACE: sampling perfection with application optimized contrasts using different flip angle evolution
FSL: FMRIB Software Library

